# Genomic epidemiology of circulating SARS-CoV-2 variants during first two years of the pandemic in Colombia

**DOI:** 10.1101/2022.06.20.22275744

**Authors:** Cinthy Jimenez-Silva, Ricardo Rivero, Jordan Douglas, Remco Bouckaert, Julian Villabona-Arenas, Katherine Atkins, Bertha Gastelbondo, Alfonso Calderon, Camilo Guzman, Daniel Echeverri-De la Hoz, Marina Muñoz, Nathalia Ballesteros, Sergio Castañeda, Luz H. Patiño, Angie Ramirez, Nicolas Luna, Alberto Paniz-Mondolfi, Hector Serrano-Coll, Juan David Ramirez, Salim Mattar, Alexei Drummond

## Abstract

The emergence of highly transmissible SARS-CoV-2 variants has led to surges in cases and the need for global genomic surveillance. While some variants rapidly spread worldwide, other variants only persist nationally. There is a need for more fine-scale analysis to understand transmission dynamics at a country scale. For instance, the Mu variant of interest, also known as lineage B.1.621, was first detected in Colombia and was responsible for a large local wave but only a few sporadic cases elsewhere. To provide a better understanding of the epidemiology of SARS-Cov-2 variants in Colombia, we used 14,049 complete SARS-CoV-2 genomes from the 32 states of Colombia, and performed Bayesian phylodynamic analyses to estimate the time of variants introduction, their respective effective reproductive number, and effective population size, and the impact of disease control measures. We detected a total of 188 SARS-CoV-2 Pango lineages circulating in Colombia since the start of the pandemic. We showed that the effective reproduction number oscillated drastically throughout the first two years of the pandemic, with Mu showing the highest transmissibility (Re and growth rate estimation). Our results reinforce that genomic surveillance programs are essential for countries to make evidence-driven interventions towards the emergence and circulation of novel SARS-CoV-2 variants.

## Background & Summary

Colombia reported its first confirmed SARS-CoV-2 infection on 6 March 2020 in a traveler returning from Milan, Italy^1^. By April 2022, the country had reported more than 6 million SARS-CoV-2 infections and over 135,000 deaths^2^ (Figure 1). According to epidemiological data, the SARS-CoV-2 epidemic in Colombia has been characterized by four pandemic waves with exponential growth in cases^3^. A wide range of strategies has been implemented to mitigate these surges of cases. That includes restrictions on mobility (such as school and airport closures), advice on mask use and physical distancing in public places, and vaccination^4–7^. Nevertheless, the current number of cases shows that the transmission of the virus is far from being under control, and those mitigation strategies may be insufficient^8^. However, the difficulties in achieving control were unlikely caused by strategy choice but rather by changes in the virus’s transmissibility^9^.

**Figure 1.**
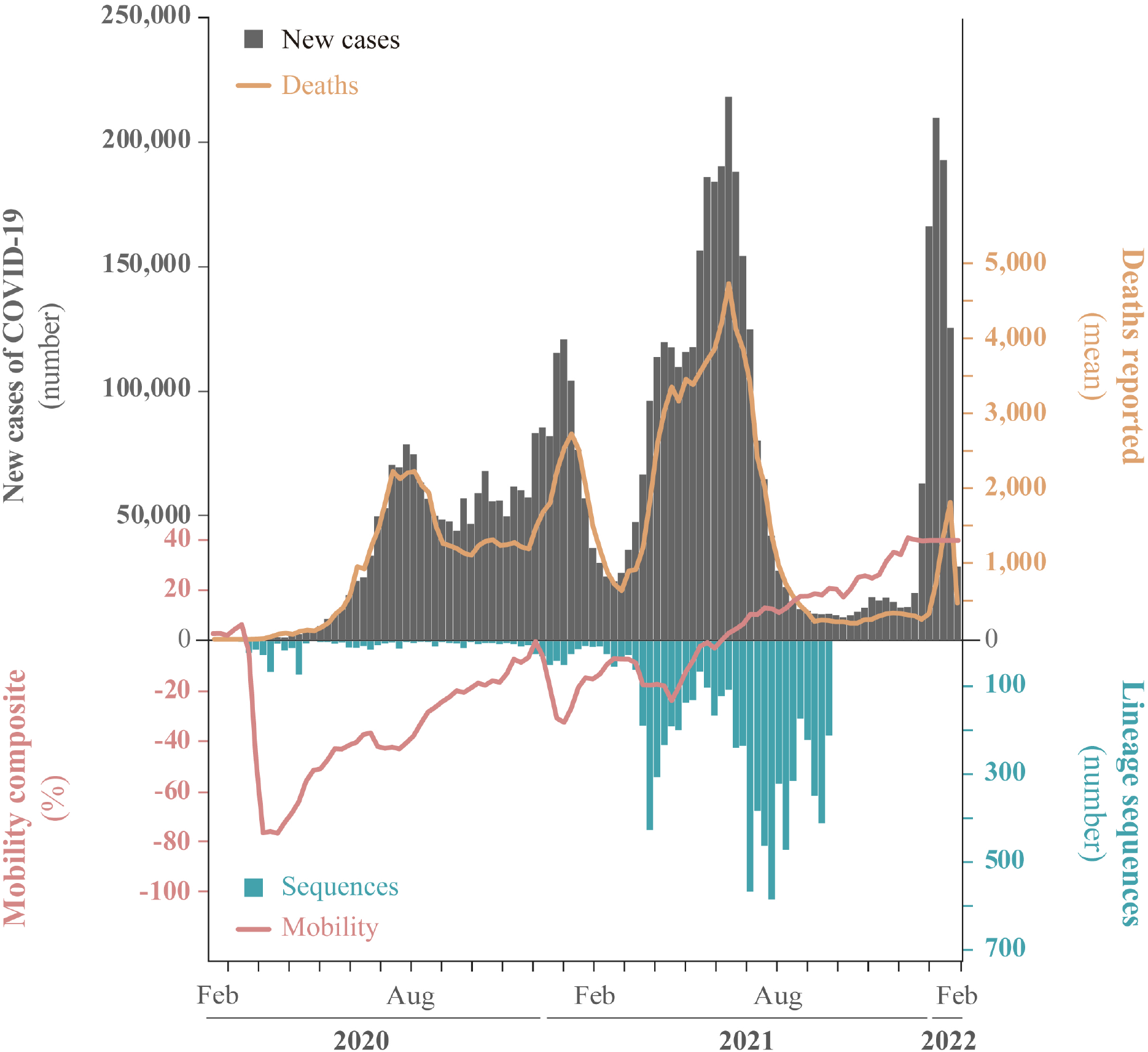
Overview of the COVID-19 confirmed cases were sampled in Colombia during the two first years of the pandemic. Top: Number of daily reported cases (gray bars) and deaths (yellow line), up until February 2022. Bottom: total number of sequences (blue bars) and mobility data from 32 states in Colombia (red line) taken from covid19.healthdata.org.

SARS-CoV-2 genetic diversity has been described using lineages, and a multiple nomenclature system has been established ^10^. Notably, large-scale sequencing has led to the identification of genetic variations with enhanced transmissibility, virulence, or evasion of host immune response around the world ^11,12^. The Technical Advisory Group on Virus Evolution from World Health Organization (WHO) labels the variants that pose an increased risk to global public health ^13^ as ‘Variants of Concern’ (VOC). These include Alpha (WHO nomenclature) or B.1.1.7 (Pango nomenclature), Beta (B.1.351), Gamma (P.1), Delta (B.1.617.2 + AY.x), and Omicron (BA.1 BA.2). Genomic surveillance has also led to identifying variants that carry mutations in the spike protein that may confer higher transmissibility and immune escape (such as mutations D614G, E484K/Q, K417T, N501Y, and P681H) ^14–17^. These variants are termed ‘Variants of Interest’ (VOI) and include Mu (B.1.621) and Lambda (C.37) ^14, 18–21^. Moreover, genomic surveillance has enabled phylodynamic investigation that has been vital to understanding global and local dynamics and tracing the zoonotic and time of origins ^15,22,23^.

After its first report in September 2020, the alpha variant soon spread around the world, and became the dominant variant in many countries. But this was not the case in Colombia, where other important variants might have been circulating instead. For instance, the VOI Mu was first detected in Colombia and was responsible for large local outbreaks (in the presence of other VOCIs, including Alpha) but caused a few sporadic outbreaks elsewhere. Phylodynamics analysis of SARS-CoV-2 has led to many significant findings, but more insights are needed from different epidemiological settings to understand better its spread and more effective approaches to control. To help achieve this, we aim to describe epidemiological trends and characterize the spatio-temporal dynamics of the most prevalent SARS-CoV-2 variants in Colombia using Bayesian Coalescent Skyline and Birth-death Skyline phylodynamic models. This study is the first recompilation that describes the epidemiological trends and circulating SARS-CoV-2 variants during the first two years of the pandemic in Colombia from a phylodynamic perspective.

## Results

The first documented case of SARS-CoV-2 infection in Colombia was reported on 3 March 2020. Initially, molecular testing and genome sequencing of SARS-CoV-2 were only performed by the National Health institute (INS), but capacity was increased with an additional 21 sequencing laboratories serving the 32 Colombian states ^24^.

As of February 2022, genomics surveillance in Colombia has generated 14,049 SARS-CoV-2 complete genomes, which represent 0.2% of the 5.9 million confirmed cases during this period. Compared with other South American countries, Colombia generated the fifth highest volume of SARS-CoV-2 genomic data during the first two years of the COVID-19 pandemic (Table 1). We generated 610 novel sequences from three different Colombian states for this study, and 13,444 were downloaded from GISAID.

**Table 1.** SARS-CoV-2 sequences available in GISAID from each country of South America until 18th February 2022 vs. number of cases (https://www.statista.com/statistics/1101643/latin-america-caribbean-coronavirus-cases/).

| Country | Total of Sequences | Number of cases |
| --- | --- | --- |
| Brazil | 114,181(0.40%) | 27,940,119 |
| Chile | 21,539(7.83%) | 2,747,55 |
| Argentina | 16,501(0.18%) | 8,799,858 |
| Peru | 15,736(0.45%) | 3,474,965 |
| Colombia | 14,653(0.24%) | 6,035,143 |
| Ecuador | 4,299(0.53%) | 808,925 |
| Paraguay | 1,215(0.19%) | 632,444 |
| Suriname | 1,027(1.32%) | 77,549 |
| Uruguay | 743(0.09%) | 800,833 |
| Venezuela | 297(0.05%) | 508,968 |
| Bolivia | 236(0.02%) | 887,089 |
| Guyana | 63(0.10%) | 62,537 |

We reconstructed the dynamics of the 10 predominant SARS-CoV-2 variants that circulated in Colombia using Bayesian phylodynamic modeling. These methods allowed us to estimate each variant’s transmissibility with, effective reproductive number (*R*_*e*_) and effective population size (*N*_*e*_) (further details about this analysis are in the methods section).

### Variant classification and distribution

The 14,049 SARS-CoV-2 complete genomes were grouped in 188 SARS-CoV-2 Pango lineages, which have circulated in Colombia. Despite the vast genetic diversity documented, only ten SARS-CoV-2 variants were predominant during the two first years of the pandemic (Figure 2). Herein, we use the pango nomenclature name for those variants that are not labeled as VOIC (variants of interest or concern). These ten lineages include four pango lineages (B.1, B.1.1.1, B.1.420, and B.1.1.348), four variants of concern (Alpha: B.1.1.7; Gamma: P.1; Delta: B.1.617, AY.x; and Omicron: B.1.1.529, BA.1, BA.2.x), and two variants of interest (Lambda: C.37; and Mu: B.1.621, BB.1 and BB.2) ^13^. The most populated states of Colombia (Antioquia, Cundinamarca, and Valle del Cauca) generated the highest number of sequences (greater than 2000 sequences each). Mu, Delta, and Gamma variants were documented in 31, 28 and 29 states (out of 32). Alpha and Lambda were documented in 12 states, and Omicron was documented in 17 states. The most widespread variant, Mu, showed the highest prevalence in the capital district (Bogotá) (19.43%), and Antioquia state (19%)(Figure2B, Figure2C).

**Figure 2.**
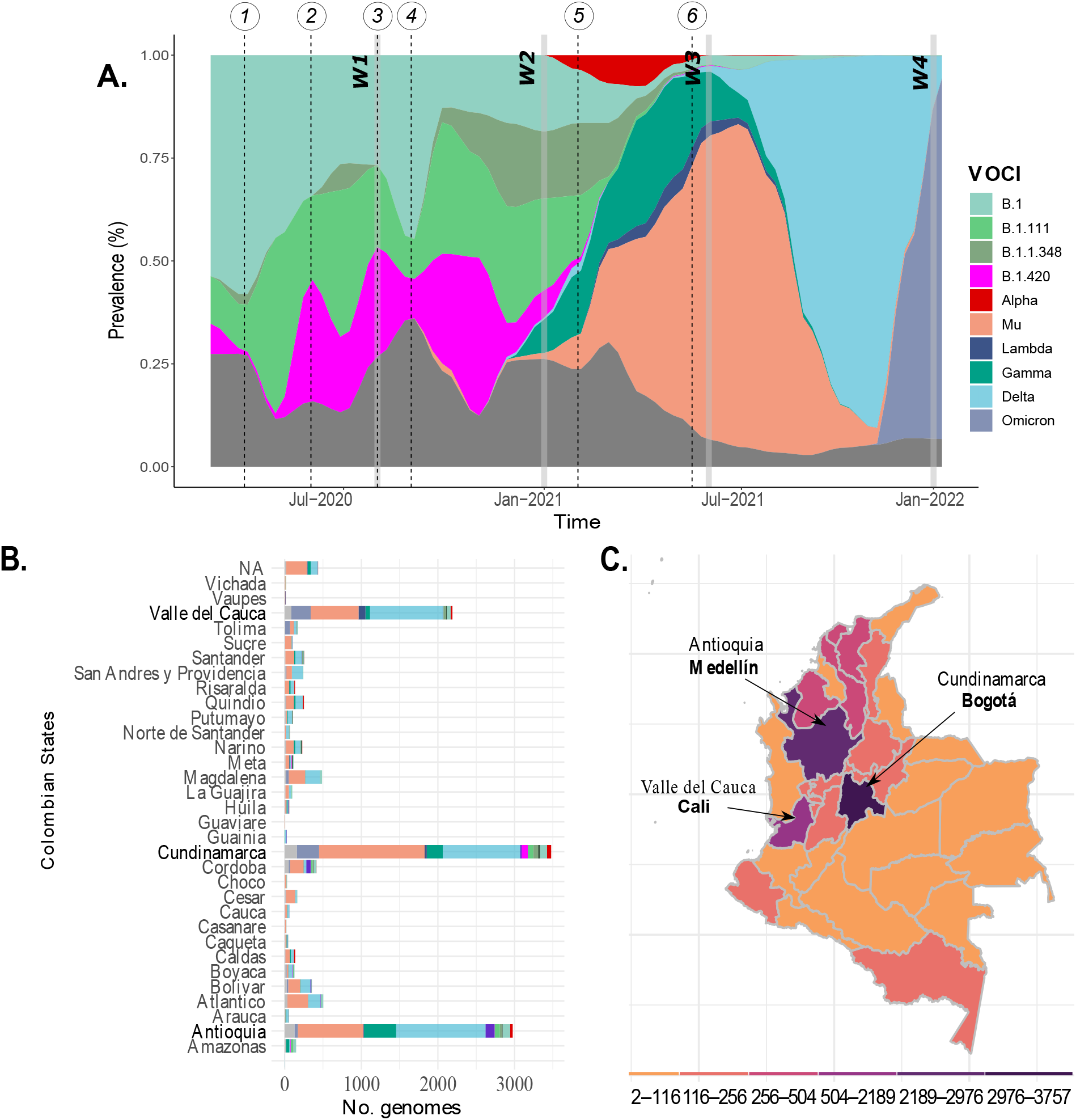
SARS-CoV-2 genetic diversity. A) shows the most prevalent Pangolin COVID-19 global lineages, SARS-CoV-2 Variants of Interest and SARS-CoV-2 variant of concern (VOCIs) until February 2022 in Colombia. Horizontal grey bars represent the four waves (W1, 2, 3, 4). The black dotted line indicated 1) the first National Lockdown in Colombia on March 25, 2020, 2) end of the first National lockdown on 18 June 2020, decreasing the restriction measures for controlling COVID-19, 3) Policies implementation for economic reactivation 10 August 2020, 4) Reopening of domestic and international flights during September 2020, 5) Vaccination phase 1 implemented on 17 February 2021, and 6) Vaccination phase 2 implemented on 17 June 2021. B) indicates the number of COVID-19 genomes isolated from 32 states of Colombia, and the capital city (Bogotá) available in GISAID and the sequences obtained in this study. C) Colombian map indicating the total genome collected per state.

In the following sections, we describe the dynamics of COVID-19 in terms of the four COVID-19 waves reported in Colombia and considering the dates on which specific Colombian measures and strategies were raised and implemented, as was used by ^4^ which defined the first two pandemic periods in Colombia:

### First period of the pandemic: from 26 February to 10 August 2020

The first wave of the COVID-19 pandemic in Colombia ran from when the first case was reported (26 February 2020) until just before the subsequent relaxation of the stringent non-pharmaceutical interventions (NPIs) that were implemented (06 March 2020). These NPIs included the declaration of a national emergency, closing of schools and universities, restriction of international flights, the closing of the international borders, and the first lockdown (Between 25 March 2020 and 18 June 2020). 552,523 cases were reported during this wave, 408 (0.07%) were sequenced, and 22 variants were identified. B.1 was the predominant (46.3%) variant, followed by B.1.111 (21.8%), B.1.420 (9.8%), and B.1.1.348 (1.71%), which co-circulated after their emergence. We dated the most recent common ancestors to exist around the 23 February 2020 (95% credible interval (CI) of 28 November 2019 to 28 February 2020), 27 February 2020 (CI: 10 February 2020 - 14 March 2020), 10 April 2020 (CI: 26 February 2020 - 24 March 2020), and 28 March 2020 (CI: 02 March 2020 - 20 April 2020) for B.1, B.1.111, B.1.420, and B.1.1.348, respectively (Figure 3). This suggests that SARS-COV-2 could have circulated earlier than first reported, but there is considerable uncertainty around the precise date. Values of Re ranged between 0.37 and 1.88 for B.1, between 0.26 and 3.49 for B.1.111, between 0.47 and 2.08 for B.1.420, and between 0.47 and 2.54 for B.1.1.348 (Supplementary Fig.1). The values of *R*_*e*_ remained relatively constant, with an average value of around 0.92 and 1.33 for variants B.1 and B.1.1.348, respectively. In contrast, there were two peaks with values greater than 1.5 for variants B.1.420, B.1.1.348, and B.1.111. After their emergence, the population size (*N*_*e*_) rapidly increased for all these variants. *N*_*e*_ reached a plateau for all variants and remained constant for variants B.1, B.111, and B.1.420. Still, it decreased for variant B.1.1.348 after experiencing some oscillations (Supplementary Fig.2). We observed a significant(p<0.05) positive correlation between *R*_*e*_ changes and the stringency index for the variants B.1.111 and a positive correlation between differences in the effective population sizes (*N*_*e*_) and mobility changes for the variants B.1, B.1.111, B.1.1.348, and B.1.420.

**Figure 3.**
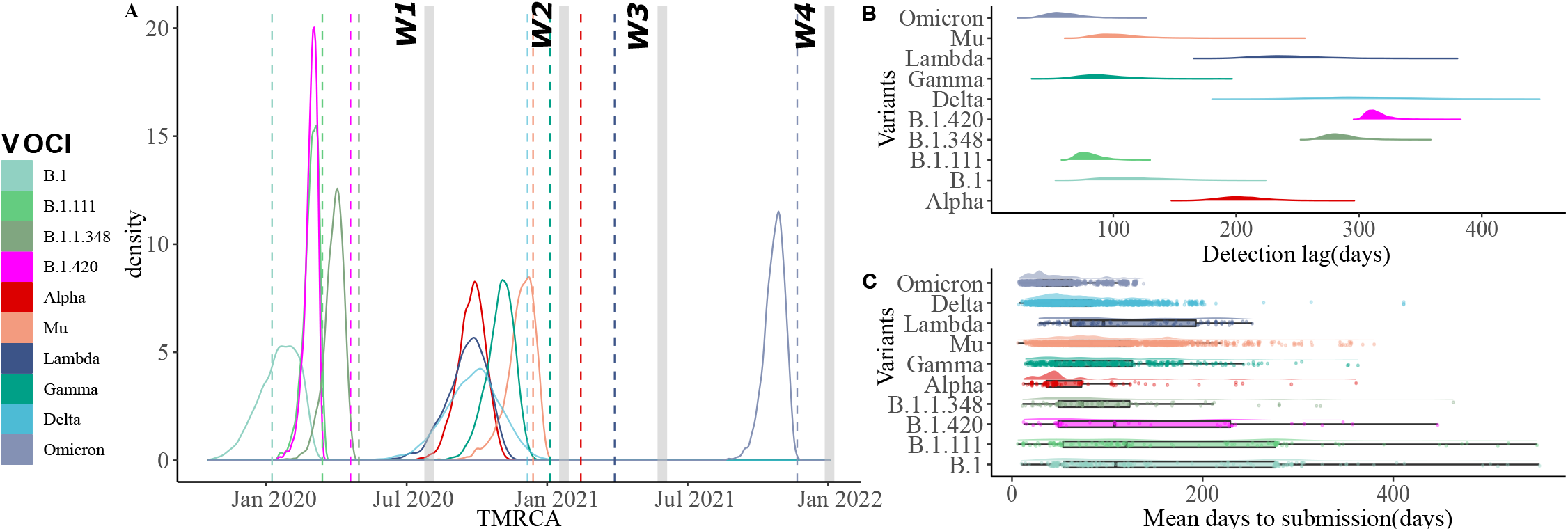
Temporal dynamics of the most prevalent variants (VOCIs). A) Time of the Most Common Ancestor of the variants that have circulated in Colombia. Horizontal grey bars represent the four waves (W1, 2, 3, 4). The dotted line indicated the first Colombian report for each variant. B) Detection lag over time as a function of TMRCA for each variant given by days. C) Mean days to submission per variant and variant as a function of submission date versus collection date.

**Figure 4.**
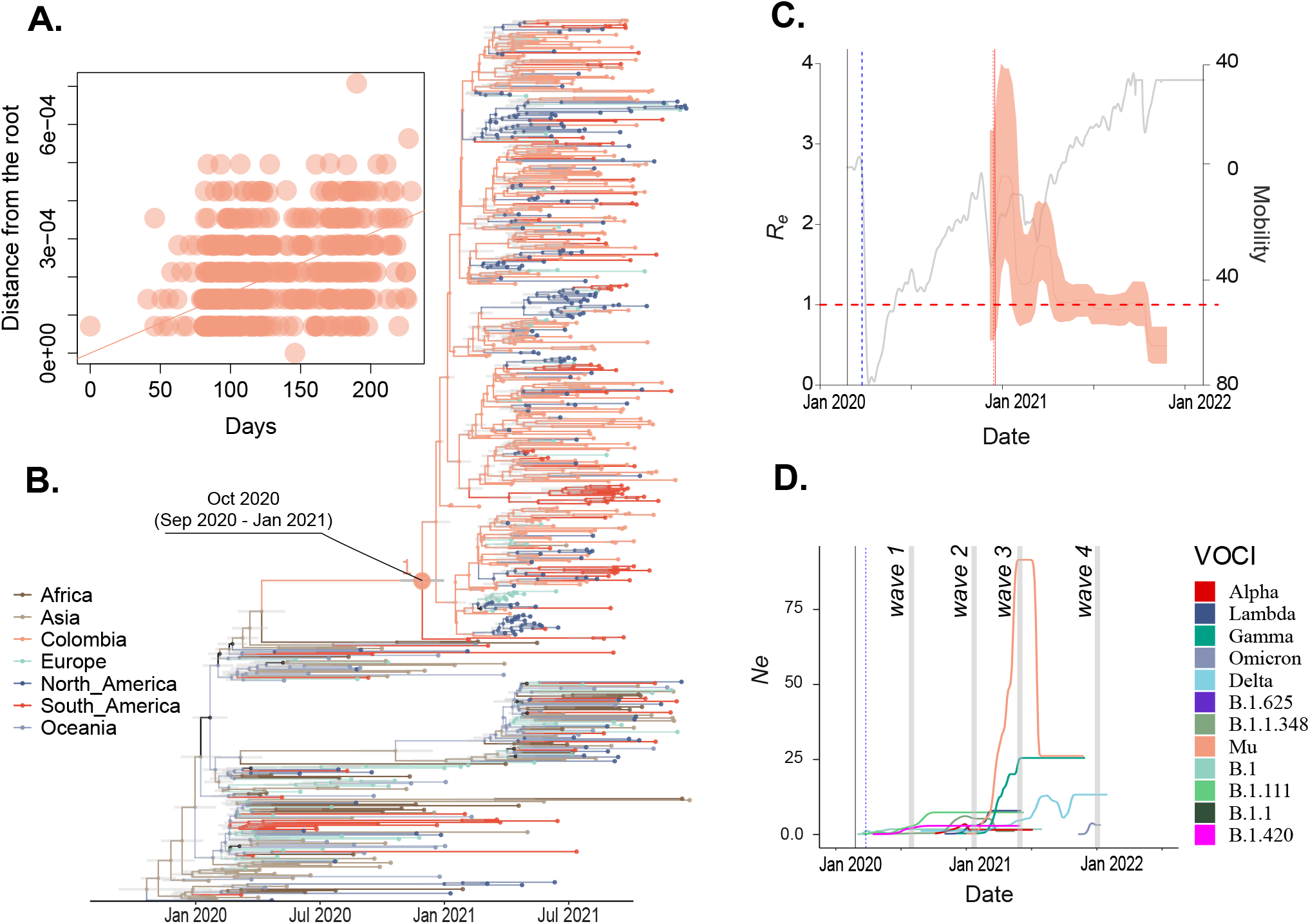
Phylodynamic of Mu variant population circulated in Colombia. A) Clock signal in Colombian sequences of the Mu variant. B) MCC tree pointing out the Mu variant origin. C) Effective reproductive number (*R*_*e*_) of Mu and D) Effective population size (*N*_*e*_) of all variants in Colombia over time. The phylogenetic tree was built with all the major Global Outbreak variants with the most representation on B.1.621 lineage (Mu variant). Branches’ colors represent Global regions and Colombia (country).

### Second period of the pandemic: from 10 August 2020 to 6 March 2021

The Colombian government implemented decreased the domestic response measures for controlling COVID-19 on 10 August, and simultaneously promoted a COVID-19 testing program, contact tracing, and sustainable selective isolation ^25^. With COVID-19 measures relaxing across Colombia (which included lifting mobility restrictions and opening domestic travel), the country experienced a new surge of cases, 2,499,104 cases were reported during this wave, 668 (0.034%) were sequenced, and 51 variants were identified. The variants B.1.111, B.1, B.1.1.348 and B.1.420 continue to predominate during this period and represented respectively 21.4%, 17.5%, 14.8% and 11.3% of the cases. Values of *R*_*e*_ ranged between 0.21 and 1.37 for B.1.111, between 0.39 and 2.04 for B.1, between 0.31 and 2.01 for B.1.1.348, and between 0.47 and 2.54 for B.1.420 (Supplementary Fig.1). The values of *R*_*e*_ remained relatively constant with an average value of around 0.91, 1.08, and 1.08 for variants B.1.111, B.1, and B.1.1.348, respectively. During this period, the Technical Advisory Group on Virus Evolution from World Health Organization (WHO) introduced the notion of ‘Variants of Concern’ (VOC) and ‘Variants of Interest’ (VOI) ^13^. During this period in Colombia, three variants of concern were reported, Gamma (5% of the cases), Delta (0.5%) and Alpha (0.59%), and one variant of interest, Mu (2.9%). All these variants co-circulated after their emergence, and the most recent common ancestors of the variants documented in Colombia were dated to 23 August 2020 (CI: 18 July 2020 - 02 October 2020), 01 December 2020 (CI: 25 September 2020 - 06 December 2020), 09 December 2020 (CI: 27 October 2020 - 09 January 2021), and 30 September 2020 (CI: 27 August 2020 - 11 October 2020) for Delta, Gamma, Mu, and Alpha, respectively (Figure 3). Compared with the first report for each variant, this suggests that Delta and Gamma could have circulated weeks earlier than the first report. Still, there is considerable uncertainty around the precise date. Although the Lambda variant was not reported during this period, the TMRCA was estimated around 15 December 2020 (CI: 22 October 2020 - 01 February 2021) (Figure 3). The values of *R*_*e*_ ranged between 0.45 and 2.68 for Delta, between 0.40 and 2.78 for Gamma, between 0.73 and 4.0 for Mu, and between 0.28 and 2.11 for Alpha (Supplementary Fig.1). The values of *R*_*e*_ remained relatively constant with an average value of around 0.91, 1.08, and 1.08 for variants B.1.111, B.1, and B.1.1.348, respectively. Concerning the VOCIs circulated during this period, values of *R*_*e*_ ranged between 0.45 and 2.68 for Delta, between 0.40 and 2.78 for Gamma, between 0.73 and 4.0 for Mu, and between 0.28 and 2.11 for Alpha (Supplementary Fig.1). The values of *R*_*e*_ remained relatively constant with an average value of around 1.29, 1.24, 1.8, and 1.20 for variants Delta, Gamma, Mu, and Alpha, respectively. The population size (*N*_*e*_) showed a clear upward trend before the highest peak during this period for Lambda and Alpha variants after their emergence. This was followed by a plateau and remained constant for both variants. Although Gamma, Mu, and Delta were reported during this period, *N*_*e*_ reached low values and increased rapidly before waves 3 and 4, respectively (Supplementary Fig.2). We observed a significant (<0.05) positive correlation between *R*_*e*_ changes and three or four predictors evaluated (mobility, vaccinated people, and stringency index for Delta, Gamma and Mu; and mask use for Alpha as well). However, the highest R-squared values were for mobility (p> 0.5) for Delta, Gamma, and Alpha variants. There is a positive correlation between differences in the effective population sizes (*N*_*e*_) and mobility changes for the variants Delta, Gamma, and Alpha.

### Third period of the pandemic: from 7 March 2021 to July 2021

During the third wave, Colombia reported 1,797,454 cases, higher than previous waves. 2,984 samples (0.16%) were sequenced, and 44 variants were identified. The most prevalent variant during this time was Mu, with 48.42% of the cases. Gamma, Lambda, and Alpha were predominated representing (21.41%), (4.59%), and (4.79% of the cases), respectively. Delta was documented in low proportion (0.33%). Values of *R*_*e*_ ranged between 0.64 and 2.76 for Gamma, between 0.59 and 2.86 for Lambda, between 0.25 and 1.60 for Alpha, between 0.75 and 2.28 for Mu, and between 0.56 and 2.32 for Delta (Supplementary Fig.1). The values of *R*_*e*_ remained relatively constant with an average value of around 1.52, 1.23, 0.82, 1.17, and 1.37 for all variants previously mentioned. We observed the highest peak of the population size (*N*_*e*_) for Mu and Gamma variants after their emergence. This was followed by a significant decrease and remained constant for both variants. Concerning to Delta variant, *N*_*e*_ increased rapidly after waves 3 and 4 (Supplementary Fig.2). The growth advantage dynamic values evidenced that Mu variant had an advantage over Lambda, Gamma, Alpha, and Delta during wave 3 (Supplementary Fig.3). We observed a significant (p<0.05) positive correlation between *R*_*e*_ changes and four predictors evaluated (mobility, stringency index, vaccinated people, and mask use) for the Mu variant and vaccinated people for the Lambda variant, being this predictor the highest R-squared values with 0.49 and 0.62, respectively. There is a positive correlation between differences in the effective population sizes (*N*_*e*_) and the Number of case changes for the Mu variant. Still, there was no significant association between *R*_*e*_ and mobility changes for the Mu variant during this wave (Tables 2 and 3).

**Table 2.** Relationships between possible predictors of the effective reproductive number of the most predominant variants of COVID-19. We tested five predictors using a linear regression between a response variable (*R*_*e*_ of each variant) and one variable or predictor (explanatory variables). The values in the table show the adjusted R-squared and (p-value). *: statistically significant. NA means that the variable was not recorded for all period of time that a particular variant circulated.

| Lineage | mobility | stringency index | vaccinated | mask use | cases |
| --- | --- | --- | --- | --- | --- |
| B.1.1 | 0.03(0.05) | -0.0039(0.39) | NA | -0.011(0.64) | -0.01(0.92) |
| B.1.111 | -0.01(0.63) | 0.4(1.105e-08)* | NA | 0.53(3.223e-12)* | -0.008(0.49) |
| B.1.1.348 | 0.019(0.15) | -0.016(0.70) | NA | 0.2(8.88e-05)* | 0.28(1.98e-05)* |
| B.1.420 | 0.20(0.045) | -0.026(0.44) | NA | -0.044(0.56) | 0.53(0.0007)* |
| B.1.617+AY (Delta) | 0.54(< 2.2e-11)* | 0.40(< 3.82e-08)* | 0.36(< 2.62e-05)* | 0.47(< 1.67e-09)* | -0.017(0.91) |
| P.1 (Gamma) | 0.51(8.17e-09)* | 0.504(1.33e-08)* | 0.57(2.44e-09)* | 0.50(1.5e-08)* | 0.091(0.021) |
| B.1.1.7 (Alpha) | -0.001(0.33) | -0.01(0.51) | -0.02(0.54) | 0.25(0.0025)* | 0.153(0.01) |
| B.1.621 (Mu) | 0.493(2.21e-08)* | 0.252(0.00019)* | 0.498(1.80e-08)* | 0.46(8.921e-08)* | 0.012(0.21) |
| C.37 (Lambda) | -0.047(0.84) | 0.03(0.21) | 0.62(0.00029)* | 0.15(0.039) | 0.753(1.002e-07)* |
| Omicron | 0.1(0.14) | 0.48(0.006) | 0.39(0.017) | 0.16(0.101) | 0.79(5.89e-05)* |

**Table 3.** Relationships between possible predictors of the effective population sizes of the most predominant variants of COVID-19. We tested five predictors using a linear regression between a response variable (*N*_*e*_ of each variant) and one variable or predictor (explanatory variables). The values in the table show the adjusted R-squared and (p-value). *: statistically significant. NA means that the variable was not recorded for all period of time that a particular variant circulated.

| Lineage | mobility | stringency index | vaccinated | mask use | cases |
| --- | --- | --- | --- | --- | --- |
| B.1.1 | 0.11(0.00042)* | 0.14(0.000044)* | NA | 0.60(< 2.2e-16)* | 0.11(0.00035)* |
| B.1.111 | 0.11(< 2.2e-16)* | 0.25(5.648e-08)* | NA | 0.21(9.996e-07)* | 0.43(6.627e-14)* |
| B.1.1.348 | 0.37(9.939e-12)* | 0.19(2.198e-06)* | 0.82(< 2.2e-16)* | 0.02(0.08) | 0.08(0.0018) |
| B.1.420 | 0.72(< 2.2e-16)* | 0.36(2.182e-11)* | NA | 0.27(1.006e-08)* | 0.47(1.197e-15) |
| B.1.617+AY (Delta) | 0.54(2.26e-11)* | 0.40(3.82e-08)* | 0.36(2.621e-05)* | 0.47(1.67e-09)* | -0.017(0.910) |
| P.1 (Gamma) | 0.51(< 2.2e-16)* | 0.54(< 2.2e-16)* | 0.60(< 2.2e-16)* | 0.49(< 2.2e-16) | -0.009(0.72) |
| B.1.621 (Mu) | -0.0006(0.33) | 0.029(0.04) | -0.009(0.84) | -0.008(0.65) | 0.51(< 2.2e-16)* |
| B.1.1.7 (Alpha) | 0.10(0.00071)* | 0.083(0.0021) | 0.08(0.007) | -0.01(0.92) | -0.006(0.521) |
| C.37 (Lambda) | -0.007(0.60) | 0.21(6.51e-07)* | 0.481(1.587e-13)* | 0.41(2.262e-13)* | 0.051(0.013) |
| Omicron | 0.11(0.00042)* | 0.14(4.393e-05)* | 0.87(< 2.2e-16)* | 0.61(< 2.2e-16)* | 0.11(0.0004)* |

### Fourth period of the pandemic: from August 2021 to February 2022

This period was characterized by a significant decrease at the end of 2021 and an exponential increase in the number of cases called wave 4. During this period was, reported 1,769,695 cases, 9,946 samples (0.56%) were sequenced, and 41 variants were identified, including 83 Delta AY.x sub-variants and two Omicron, Mu, and Lambda sub-variants. As of February 2022, Omicron is the last variant of concern globally reported. In Colombia, Omicron displaced the previous variants described with a predominance of 87.6% in two first months in 2022 and has been detected in all the 32 states of Colombia. Since its identification, its prevalence has been 12% of the total variant samples identified in Colombia. The estimation of the TMRCA suggested that Omicron was introduced on 24 October 2021 (CI: 25 September 2021 - 15 November 2021) (Figure 3), which is congruent with the first epidemiological report. Delta prevalence was 5.3%, and other variants were 7% during the same period. Values of *R*_*e*_ ranged between 0.68 and 1.79 for Delta, between 0.26 and 1.29 for Mu, and between 0.45 and 3.46 for Omicron (Supplementary Fig.1). *N*_*e*_ values increased rapidly before the highest peak of wave four, followed by a plateau, while *N*_*e*_ values maintained constant for Delta and Mu variants for this last period (Supplementary Fig.2). The highest average value of *R*_*e*_ was 1.47 for Omicron compared to 1.09 and 0.82 for Delta and Mu, respectively, during this period. The growth advantage dynamic values showed the same trend as the *R*_*e*_ values, which omicron variant evidenced an advantage compared to Delta and Mu variant during wave 4 (Supplementary Fig.3). A significant (p<0.05) positive correlation between *R*_*e*_ changes and the number of cases was significant, while between *N*_*e*_ changes and all five predictors evaluated were significant, with the highest R-squared for vaccinated people with 0.61 (Tables 2 and 3).

## Discussion

The present study provides a comprehensive description of the emergence and dynamic of SARS-CoV-2 variants in Colombia based on genomic surveillance and a phylodynamic approach. Variant diversity in Colombia was characterized by multiple SARS-CoV-2 variants and multiple introductions derived from ancestral B.1 lineage, which was imported mainly from European countries (Spain and Italy)^26,27^. Despite documenting at least 188 lineages in the community, only ten dominated, suggesting high transmissibility of those variants of interest and concern compared with other emerged variants. Thus, we were interested in comparing transmission differences between Colombia’s main circulated SARS-CoV-2 variants.

We employed Bayesian phylodynamic methods to recover Colombia’s most prevalent genetic variants following the first reported case in March 2020^1^, which led to COVID-19 being declared a health emergency one week later. Different control strategies have been implemented since then, such as mandatory isolation, epidemiological follow-up for air passengers who arrived in Colombia with COVID-19 symptoms, the closing of borders, and international air travel being banned on 20 March 2020^28^. Contention measures were taken on 25 March when lockdown and domestic air-travel ban was decreed, except for essential workers (such as bank tellers, post officers, and healthcare professionals)^29^.

Despite the contention measures and an observed 80% reduction in mobility, cases continued to surge throughout the country (Figure 1). The effective reproductive number (*R*_*e*_) was shown with particular fluctuation through time per each variant circulating in the country. All analyzed variants recovered peaks >1, suggesting that a variant’s spread was greater than another during a specific period (Figure S4). Comparing variants, Mu showed the highest *R*_*e*_ values indicating higher transmissibility than Alpha, Gamma, Lambda, and even Delta, which had been reported as dominant variants in the countries they have circulated^30^. This suggests that once Mu emerged in Colombia, it out-competed the other variants and became the dominant one. In late 2021, Mu was eventually out-competed by Delta.

Differences in transmissibility between variants could be explained by partial immune evasion. It has been reported that VOCIs that carry K417N, E484K, and N501Y have a higher affinity towards the hACE2 receptor and enhanced immune escape abilities as observed with Gamma and Alpha in Brazil and the United Kingdom, respectively^31,32^. Late 2020 and early 2021 were characterized by the emergence of variants exhibiting advantage-conferring mutations, and despite Alpha’s increased transmissibility and innate immune escape ability (represented in mutations N501Y and 69-70)^17,32^ it did not manage to establish as the dominant variant after its introduction around 26 November 2020 and was shortly displaced by Gamma and Mu. A different scenario occurred with highly evasive variants such as Gamma, Lambda, and Mu which dominated the transmission dynamics during Colombia’s third pandemic wave.

Gamma was first detected in Manaus, a city in the Brazilian Amazon state, and has been determined to have emerged around November of 2020 as a result of an accelerated evolutionary rate of locally circulating clades. Due to its increased viral load, it rapidly spread throughout Brazil^31,33^. This variant was first detected in Colombia on 4 January 2021. Based on TMRCA estimation, we suggest that it could have been circulating in the Colombian Amazonian region by December of 2020 (Figure 3) before its introduction into 29 states.

However, the emergence of the Mu variant in Colombia caused a displacement of other variants whose circulation had previously been characterized by geographic heterogeneity, with the Pacific region (Valle del Cauca, Cauca, Nariño, and Choco states) being dominated by Lambda. In contrast, Andean states (Huila, Risaralda, Quindio, Tolima, Cundinamarca, Boyaca, Santander, Norte de Santander and Antioquia) and Amazonian states (Putumayo, Caqueta, Guaviare, Guania, Vaupes and Amazonas) had a high circulation of Gamma (Figure 2). Our findings can be explained by Mu’s high *R*_*e*_ and its partial immune escape. Mu variant is 10.6 and 9.1 times more resistant to convalescent, and BNT162b2-immunized patient sera^34,35^. Previous studies on the impact of enhanced transmissibility and partial variant immune escape have demonstrated that epidemic sizes become larger after the introduction of a highly transmissible and immune-evasive variant. It happened commonly in a scenario comprised of slow vaccine rollout and depletion of NPIs. Furthermore, the partial immune evasion of Mu could account for reinfections and breakthroughs among previously highly immune populations^36^. These data support that Mu’s higher *R*_*e*_ as described in our study (Supplementary Fig.1.) and its ability to partially escape antibody-mediated neutralization might account for Colombia’s third wave of COVID-19 cases^37^.

On the other hand, our results suggest the opposite phenomenon occurred with Delta. Once it was introduced to Colombia on 3 April 2021, it remained undetected until 10 May 2021, coinciding with Mu’s establishment and expansion. Delta prevalence increased after July of 2021 with a steady increment in the share of reported variants (Supplementary Fig.3.C). However, cases remained low throughout July until November 2021. We propose it might be due to Delta circulating in a population with a high level of immunity elicited both by vaccination and previous exposure to Mu, which has been found to cross-neutralize Delta^38^ effectively. Despite a high Delta’s *R*_0_^39^, our findings show that its circulation in Colombia did not cause an exponential surge in cases, as reflected by its *R*_*e*_ and *N*_*e*_.

In contrast, we found the Omicron variant could be responsible for the surge in cases observed through the fourth wave after its introduction into Colombia on 24 October 2021. This is based on a marked increase of *N*_*e*_ and a steady *R*_*e*_ over 1, displacing Delta circulation in the country (Supplementary Fig.3). Our results support the predicted scenarios of introducing a highly immune evasive and highly transmissible variant in a population with high levels of immunity, with an observed out-competing of variants with high transmissibility but mild immune escape such as Delta^36,40^. The probable causes for the steep rise in Omicron’s prevalence are the control measures weakness and the 1.4-fold augment in mobility (Supplementary Fig.2.b). Although a positive correlation between mobility and Omicron’s *N*_*e*_ and *R*_*e*_ was observed, suggesting Omicron’s advantages (immune escape), it was not statistically significant. Compared with the Mu variant, the impact of Omicron on public health was considerably lower, which could be explained by Colombia’s higher vaccine coverage by the end of 2021 (62%). Therefore, even though some studies have found that population immunity wanes through time either by previous infection and vaccination and confers mild protection against reinfection and breakthrough cases by Omicron variant. Vaccination continues to effectively reduce the risk of severe disease and death as found with previous variants^8,41^.

The effective population sizes (*N*_*e*_) estimations increment of each variant precede an increment in the number of cases, followed by extensive of community transmission. The oscillations in *N*_*e*_ and *R*_*e*_ could be explained by the fluctuations in mobility and preventive and control measures applied after each reported wave. As an exploratory data analysis, a general linear regression model was evaluated to identify which actions could effectively control the variant transmission represented by *R*_*e*_ and viral population growth. We evaluated four different control strategy measures, and two variables can explain *N*_*e*_ and *R*_*e*_ values (Tables 2 and 3). In most cases, mobility showed higher values of R squared with significant values, suggesting that it affects *N*_*e*_ and *R*_*e*_. Mobility indicates the more significant potential for personal contact, which can contribute to the spread of the disease. When mobility is high, the risk of COVID-19 spread may also be increased^42,43^. However, as mobility increases, taking precautions such as getting vaccinated, Colombian COVID-19 responses such as the implementation of stringent government policies (school closures, workplace closures, cancellation of public events, restriction on public gatherings, closures of public transport, stay at home requirements, information campaigns, restrictions on international movements; and international controls), and wearing masks in public areas can all reduce the risk of disease transmission. This regression analysis had limitations, such as a small number of data points. It was performed assuming a Re value per week; there were more than 20 points in all cases. In the future, it is necessary to perform a GLM including all the proposed explanatory variants into the model and assuming more epochs.

Estimating the time of the most recent common ancestor of the viral population showed that B.1.1.348 lineage as well as Alpha and Lambda variants circulated well before its first report in Colombia denotes a detection lag suggesting cryptic transmission (Figure 3). However, most introductions were estimated to have occurred several days before the first confirmed sample. The cryptic transmission period reported in this study for Colombia has been reported for other countries^44,45^.

The applicability of Bayesian phylodynamic methods is limited considering large genomic datasets, such as that of SARS-CoV-2. We employed down-sampling strategies to address these complications, allowing us to use a representative sample of both time and geography. Furthermore, we used a novel Bayesian Integrated Coalescent Epoch PlotS (BICEPS) for efficient inference of coalescent epoch models. It integrates population size parameters and introduces a set of more powerful Markov Chain Monte Carlo (MCMC) proposals for flexing and stretching trees ^46^. The present work compared the traditional Bayesian skyline model with this novel model and found congruence in effective population size estimates (Supplementary Fig.6.) and the time to the most recent common ancestor (TMRCA) per variant estimation (Table 5) between methods. The novel implementation of tree priors and proposals allows larger genomic datasets to be analyzed for tracing an emerging virus’s spread, transmission, and population dynamics for genetic surveillance reports.

In summary, the study highlights the dynamics of the most predominant genetic variants that have been reported in Colombia in terms of transmissibility and demographic dynamic. The high transmission and effective population sizes of each variant could be explained by the increase in mobility and the reduction in the government response tracker (implementation of control measures) in Colombia. Each wave was characterized by the circulation of at least one of these prevalent variants. The emergence of the highly transmissible Mu in Colombia could explain why Delta and Alpha, which were introduced previously, did not have the same impact as in other countries such as England or Brazil. Genomic surveillance has been instrumental in informing public health response against COVID-19 in many parts of the world, including New Zealand, Australia, Iceland, and Taiwan, showing how these implementations helped to successfully control the increase of COVID-19^23^. This is made accessible by pathogen surveillance platforms such as GISAID^47^, NextStrain^48^, and Microreact^49^. Here, we have demonstrated how these technologies can inform public health response in Colombia. We advocate for the widespread adoption of such technologies in the Colombian public health infrastructure and worldwide.

## Methods

### Novel Genome sequence data

We collected Nasopharyngeal swabs from 10,674 residents from: Bogotá (the Capital District), Cali (the Capital of Valle del Cauca state) and Córdoba state (The Capital city and small towns) for testing by RT-qPCR using an in-house protocol based on the amplification of SARS-CoV-2 E gene according to WHO guidelines ^50^. We sequenced positive samples with the following data availabe: travel history(the latest country of travel), patient status (Asymptomatic, mild, severe, critic, and fatal), sample collection date, and vaccination status. Our selection criteria resulted in 610 samples: 86 samples from Córdoba, 122 from Cali, and 402 from Bogotá. We purified the ARN of SARS-CoV-2 from the selected samples using the GeneJet RNA Extraction Kit (ThermoFisher Scientific, cat no. K0732) and prepared the sequencing library following the ARTIC Network protocol^51^ and sequenced the libraries using the Oxford Nanopore MinION sequencer. Then, we processed (base-calling and demultiplexing) the raw data using Guppy v3.4.6^52^ and filtered reads by quality and length to remove short, and low-quality reads (threshold lower than 20X was assumed as N). Finally, we assembled consensus genomes following the ARTIC bioinformatics pipeline^53^. Sample collection was led by the Instituto de Investigaciones Biologicas del Tropico (IIBT) at Universidad de Córdoba and the Centro de Investigaciones en Microbiologia y Biotecnologia (CIUMBIUR) at Universidad del Rosario, which are part of the official laboratories authorized by Colombia’s Ministry of Health for testing and genomic surveillance or SARS-CoV-2. Sample collection in Córdoba was approved by the Ethics committee of Universidad de Córdoba/IIBT (Acta N° 0410-2020) in compliance with CDC’s guidelines for safe work practices in human diagnostic^54^. Sample collection in Bogotá and Cali was approved by Universidad del Rosario’s Research Ethics committee (DVO005 1550-CV1400) in compliance with Helsinki’s declaration^55^. Informed consent was obtained from all patients.

### GISAID Data curation

We retrieved all SARS-CoV-2 genome sequences from Colombia shared via GISAID (N=14,049, last accessed on 2022-02-02) and combined them with the novel genome sequences. We identified the variant of each genome sequence using the Pango nomenclature^56^. We excluded sequences with bad quality based on six different control metrics implemented in Nextclade^57^: no more than 10% ambiguous characters, no more than ten mixed sites, no more than 10% of missing data (Ns > 3000), no more than two mutation clusters, number of insertions or deletions that are not a multiple of three and number of stop codons that occur in unexpected places (2 stop codons are bad), and any outlier sequence as reported by Nextstrain^58^. We also removed sequences with incongruent lineage classification between Pangolin and Nextclade. Additional information for all sequences submitted and downloaded from GISAID is available in (Supplementary tables 1 and 2). We down-sampled the alignments by variant and homogeneously through the time (to have at least one sequence per day); any variant with greater than or equal to 100 samples was considered a major variant. This down-sampling resulted in 1662 sequences distributed in 10 different alignments (Table 4).

**Table 4.** Summary of the SARS-CoV-2 sequences’ alignment from Colombia per variant of interest assuming down-sampling criteria of at least two sequences per day. The name of each variant is defined as Pangolin lineage, and WHO nomenclature is indicated in parentheses. Height: values given in days and years in parentheses. First report: Earliest date of each lineage is reported at https://cov-lineages.org/. Omicron: B.1.1.529 + BA.1/BA.1.1 lineages

| Variant | Total-seq | Min date | Max date | days | Weeks | first report Colombia (World) |
| --- | --- | --- | --- | --- | --- | --- |
| B.1.1 | 146 | 2020-03-12 | 2022-01-03 | 510 | 72 | 2020-03-12(2020-01-08) |
| B.1.111 | 84 | 2020-03-13 | 2021-06-12 | 454 | 64 | 2020-03-13(2020-03-07) |
| B.1.1.348 | 75 | 2020-04-30 | 2021-12-31 | 384 | 54 | 2020-04-30(2020-04-30) |
| B.1.420 | 37 | 2020-03-11 | 2021-06-03 | 118 | 16 | 2020-03-11(2020-07-13) |
| B.1.1.7 (Alpha) | 80 | 2021-02-15 | 2021-10-03 | 213 | 30 | 2021-02-15(2020-09-03) |
| P.1 (Gamma) | 338 | 2021-01-04 | 2021-12-01 | 332 | 47 | 2021-01-04(2020-10-01) |
| B.1.617+AY (Delta) | 193 | 2020-12-07 | 2022-01-17 | 252 | 58 | 2020-12-07(2021-01-07) |
| B.1.621 (Mu) | 416 | 2020-10-14 | 2021-12-15 | 330 | 47 | 2020-10-14(2020-12-15) |
| C.37 (Lambda) | 133 | 2021-03-30 | 2021-09-16 | 156 | 22 | 2021-03-30(2020-07-21) |
| Omicron | 160 | 2021-12-04 | 2022-01-21 | 42 | 06 | 2021-12-04(2021-09-11) |

**Table 5.** Posterior summary of the time to the most recent common ancestor (TMRCA) showing as Tree Height parameter per variant estimated using the Birth-Death Skyline model (BDSKY), Coalescent Skyline model (SKY), and Bayesian Integrated Coalescent Epoch PlotS (BICEPS). Each value is numerical as a year.

| Variant | BDSKY | 95% HPD | BICEPS | 95% HPD | SKY | 95% HPD |
| --- | --- | --- | --- | --- | --- | --- |
| B.1.1 | 1.55 | 1.47-1.66 | 1.54 | 1.42-1.68 | 1.50 | 1.41-1.62 |
| B.1.111 | 1.28 | 1.26-1.31 | 1.28 | 1.24-1.33 | 1.26 | 1.23-1.30 |
| B.1.1.348 | 1.11 | 1.07-1.16 | 1.13 | 1.07-1.20 | 1.11 | 1.06-1.18 |
| B.1.420 | 1.09 | 1.06-1.12 | 1.16 | 1.12-1.20 | 1.13 | 1.12-1.15 |
| B.1.1.7 (Alpha) | 0.87 | 0.80-0.94 | 0.85 | 0.75-0.95 | 0.83 | 0.75-0.92 |
| P.1 (Gamma) | 1.07 | 0.9-1.2 | 1.07 | 0.93-1.39 | 1.03 | 0.9-1.13 |
| B.1.617+AY (Delta) | 1.68 | 1.51-1.86 | 1.63 | 1.43-1.83 | 1.63 | 1.43-1.85 |
| B.1.621 (Mu) | 0.72 | 0.62-0.90 | 0.66 | 0.62-0.73 | 0.70 | 0.62-0.80 |
| B.1.1.7 (Alpha) | 0.52 | 0.45-0.60 | 0.48 | 0.39-0.57 | 0.50 | 0.41-0.60 |
| C.37 (Lambda) | 0.70 | 0.60-0.80 | 0.64 | 0.52-0.77 | 0.68 | 0.56-0.82 |
| Omicron | 0.70 | 0.60-0.80 | 0.64 | 0.52-0.77 | 0.68 | 0.56-0.82 |

### Measuring the variant’s growth rate and the effective reproduction number *R*_*e*_

To estimate the growth advantage of each variant, we used the frequencies (weekly) of the SARS-CoV2 variants to fit multinomial logistic regression models that include a natural cubic spline to allow for slight variation in the growth rate of a given variant as a function of the sampling date. These multinomial spline models consider the frequencies of the major SARS-CoV2 variants as separate outcome levels (the remaining variants are aggregated in the category of ‘other variants’) to simultaneously model the competition among all variants. A model was fitted for each of the four wave periods in Colombia using the nnet package v.7.3-17 in R. v.3.5.0^59^. The models were used to produce Muller plots to display the change in the relative frequencies of the major SARS-CoV-2 variants. Furthermore, estimates of the expected multiplicative effect on *R*_*e*_ based on the relative abundance of each variant were calculated assuming a gamma distributed generation time (Mean = 4.7 days, standard deviation = 2.9) using weighted effects contrasts and the package emtrends v.1.7.3^60^,61 in R v.3.5.0 ^59^.

### Bayesian phylodynamic analysis

We aligned the sequence data of each major variant using MAFFT v 7 ^62^ and all the alignments were split by codon position. We tested the temporal signature of each alignment using a Neighbor-Joining tree that was inferred using the ape package v.5.6-2 ^63^ in R v.3.5.0 ^59^ and a regression of root-to-tip genetic distance against sampling time using TempEST v1.5.338^64^. The levels of temporal signal were assessed by visual inspection and by the correlation coefficient. All alignments showed a positive correlation (the correlation coefficient ranged between 0.0042 and 0.8) and appear to be suitable for phylogenetic molecular clock analysis in BEAST (Supplementary Fig.5). We assumed a strict molecular clock as a prior for the clock rate in all cases (a log-normal distribution with a mean of 0.001 subs/site/year and standard deviation of 0.03), assuming from previous analysis^**?**^.

We performed bayesian inference of phylogeny and estimated the time to the most recent common ancestor (TMRCA) of each node and the demographic dynamics (in terms of effective population sizes, *N*_*e*_) over time of ten different alignments (group of sequences from ten variants that were documented in Colombia) using two different tree priors: The Bayesian Coalescent Skyline (BCS) ^65^ and the recently implemented Bayesian Integrated Coalescent Epoch PlotS (BICEPS) model ^46^. We evaluated the congruence between both models and encourage to use the last one (BICEPS) because it is computationally more efficient than BCS and allows extensive data sets analysis. We estimated the effective reproduction number (*R*_*e*_) through time using a Bayesian birth-death skyline model ^66^ with ten and fourteen dimensions. Estimates of *R*_*e*_ using two different dimensions were compared to evaluate changes in the inferred *R*_*e*_ in some periods. All the posteriors values for the parameters of interest were reported as mean and credible intervals (CI), which is referred to as the 95% of high posterior density (HPD). We used the R-package bdskytools (https://github.com/laduplessis/bdskytools) to plot the smooth skyline, marginalizing our *R*_*e*_ estimates on a regular time grid (defined as the number of weeks that each variant has circulated) and calculating the HPD at each gridpoint. The models are available as packages in the platform BEAST v2.6.7 ^67^. In order to confirm the origin of Mu variant, we performed a phylogeography analysis using the Bayesian discrete phylogeography model (DPG^68^). We considered migration between seven demes (Global regions and Colombia Country) assuming that the transition rates between locations were reversible.

We determined the Hasegawa-Kishino-Yano model (HKY) to be the best-fit nucleotide substitution models ^69^ without site heterogeneity for all alignments using BModelTest v1.2.1 ^70^. We used three independent Markov Chain Monte Carlo with 400 million iterations using the CoupledMCMC package (MC3) v1.0.2 ^71^. We diagnosed the MCMC samples using Tracer v1.7.2 (http://tree.bio.ed.ac.uk/software/tracer) until they reached effective sample sizes over 200 for all parameters. We summarized Maximum clade credibility trees (MCC) using TreeAnotator package. To visualize trees and outputs, we used Figtree v1.4.4 and R v.3.5.0 ^59^ with packages: ape v5.6-2 ^72^ and ggtree v3.4.0 ^73^. All output files are available in the repository of GitHub (https://github.com/cinthylorein/Colombia-COVID-19-phylodynamics.git/).

### Metadata and Statistical Analysis

We accessed socio-demographic and COVID policy intervention variables to determine the association between each variant’s *R*_*e*_ and *N*_*e*_ using generalized linear models. The variables were change in human mobility given in % units (as measured by cell phone mobility data); vaccine coverage (shows the percentage of people who receive at least one dose of a vaccine, and those who are fully vaccinated against COVID-19); estimated infections (the number of people we estimate are infected with COVID-19 each day, including those not tested); mask use (represents the percentage of the population who say they always wear a mask in public) ^74^, and lockdown policies ^75^, which is given by the stringency index (composite measure based on nine response indicators including school closures, workplace closures, and travel bans with value from 0 to 100 = strictest). We calculated Pearson correlation coefficients to avoid excessive co-linearity among explanatory predictors removing variables that exceeded 0.7. We transformed Predictors into units space and standardized to eliminate the effect of the magnitude of different co-variants. These statistical analyses were performed using package stats v4.3.0 in R v.3.5.0 ^59^. We reported the adjusted R square and p-value of less than 0.05 were considered statistically significant.

## Data Availability

All data produced in the present study are available online at GISAID

https://github.com/cinthylorein/Colombia-COVID-19-phylodynamics.git

## Acknowledgments

This work is supported by the Marsden grant 18-UOA-096 from the Royal Society of New Zealand, Colombia’s Science Ministry (Minciencias), BPIN 20200000100090. This project was funded by the Universidad del Rosario in the framework of its strategic plan RUTA2025. Thanks to President and the University Council for leading the strategic projects (JR). We also thank the Colombian network for SARS-CoV-2 genomic surveillance led by the National Institute of Health (INS).

## Author contributions statement

C.J.S and R.R contributed to the design of the study, conceived and conducted the analysis, analysed the results and writing of the manuscript; J.D and R.B contributed to analysis of the results and writing of the manuscript; R.R, J.D.R, B.G, A.C, C.G, D.E.D, M.M, S.C and L.H.P contributed to sample collection, viral detection, clinical characterization, genome isolation and sequencing; J.D.R, M.M, S.C and L.H.P contributed to genome assembly, analysis and quality control; C.J.S, R.R, J.D.R, S.M and A.D contributed to result discussion and reviewing of the manuscript. All the authors reviewed and approved the manuscript.

## Competing interests

The authors declare no competing interests.

## Supporting Information

**Supplementary Fig.1.**
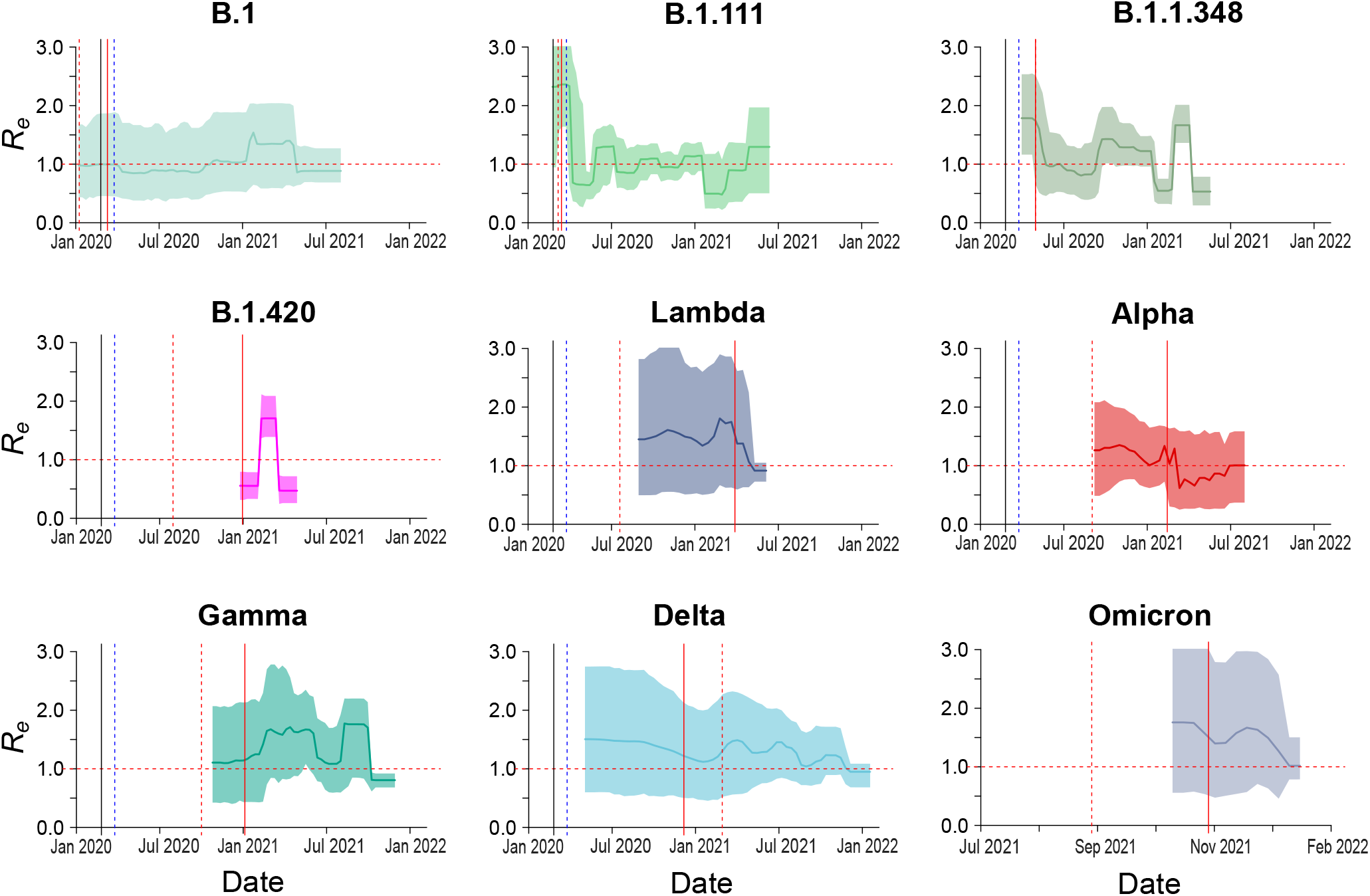
Birth-death skyline (bdsky) analysis of nine COVID-19 variants circulated in Colombia. The most prevalent Pangolin COVID-19 global lineages during the first semester of the pandemic (B.1, B.1.111, B.1.1.348, B.1.420), one designated Variants of Interest (VOIs) (Lambda) and four SARS-CoV-2 variants of concern (VOCs: Alpha, Gamma, Delta, and Omicron). The median posterior estimate of the estimated effective reproductive number (*R*_*e*_) over time is shown, with the 95% highest posterior density (HPD) interval in different colors. The horizontal red dotted line indicates the epidemic threshold (*R*_*e*_ =1). The black dotted line showed the first COVID-19 case that was identified in Colombia on February 26, 2020. The blue dotted line indicated the first National Lockdown in Colombia on March 25, 2020. The red dotted line showed the first sequence reported in https://cov-lineages.org/ for each variant. The continuous red line indicated the first Colombian sequence reported for each variant.

**Supplementary Fig.2.**
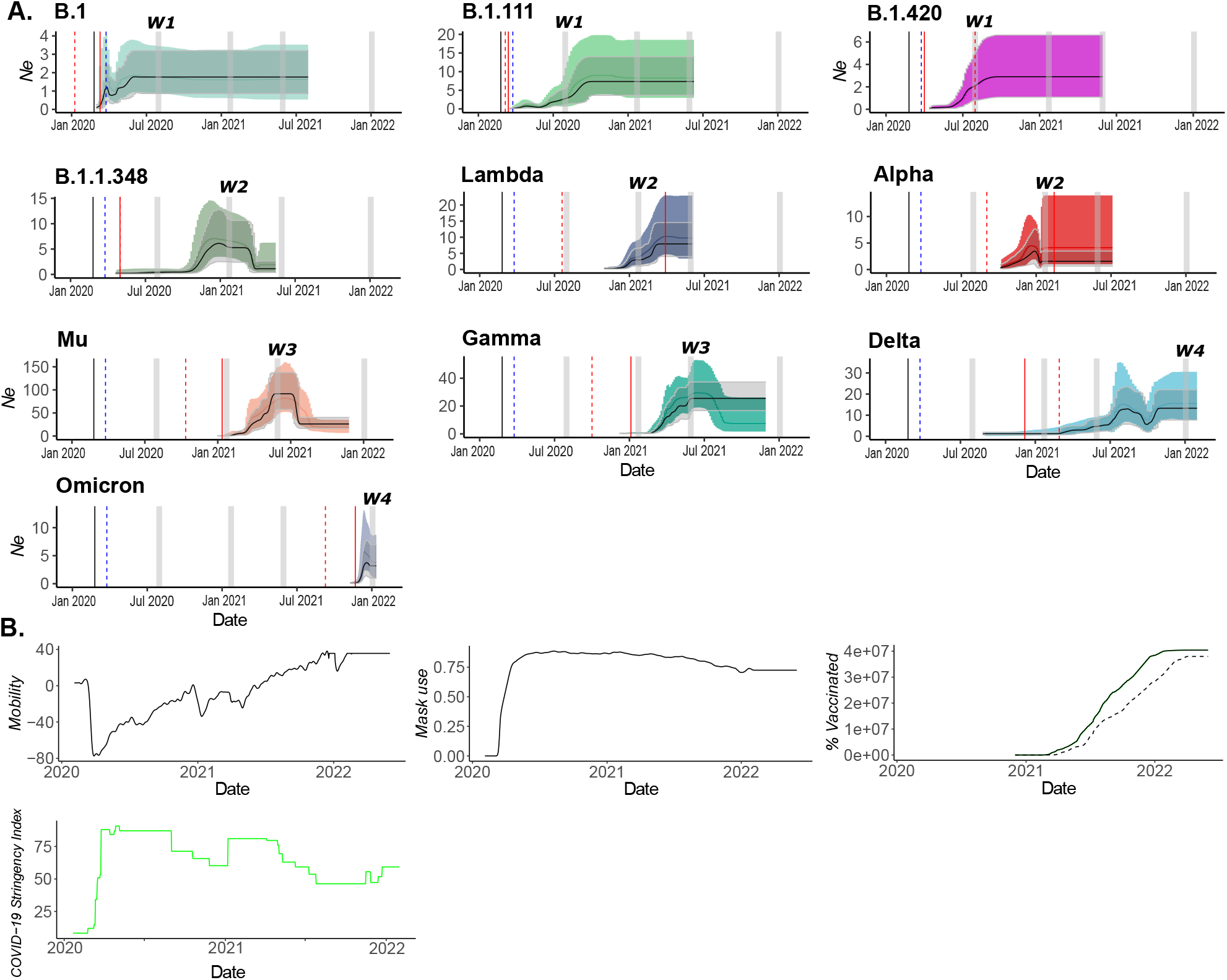
Dynamic of SARS-CoV-2 in Colombia from March 2020 to February 2022. Effective population size (*N*_*e*_) of A) The most prevalent Pangolin COVID-19 global lineages during the first semester of the pandemic (B.1, B.1.111, B.1.1.348, B.1.420), two designated Variants of Interest (VOIs) (Lambda and Mu) and four SARS-CoV-2 variants of concern (VOCs: Alpha, Gamma, Delta, and Omicron) which have circulated in Colombia using Bayesian skyline plot (in color) and BICEPS analysis (grey). Horizontal grey bars represent the four waves (W1, 2, 3, 4). *N*_*e*_ is shown on the y-axis, and the time in years is shown on the x-axis. The dark middle line indicates the median; lighter outer lines cover the 95%HPD intervals. The blue dotted line indicated the first National Lockdown in Colombia on March 25, 2020. The red dotted line showed the first sequence reported in https://cov-lineages.org/ for each variant. The continuous red line indicated the first Colombian sequence reported for each variant. B) Variables that could affect SARS-CoV-2 transmission (Mobility, Mask use, % vaccinated people, and COVID-19 Stringency index).

**Supplementary Fig.3.**
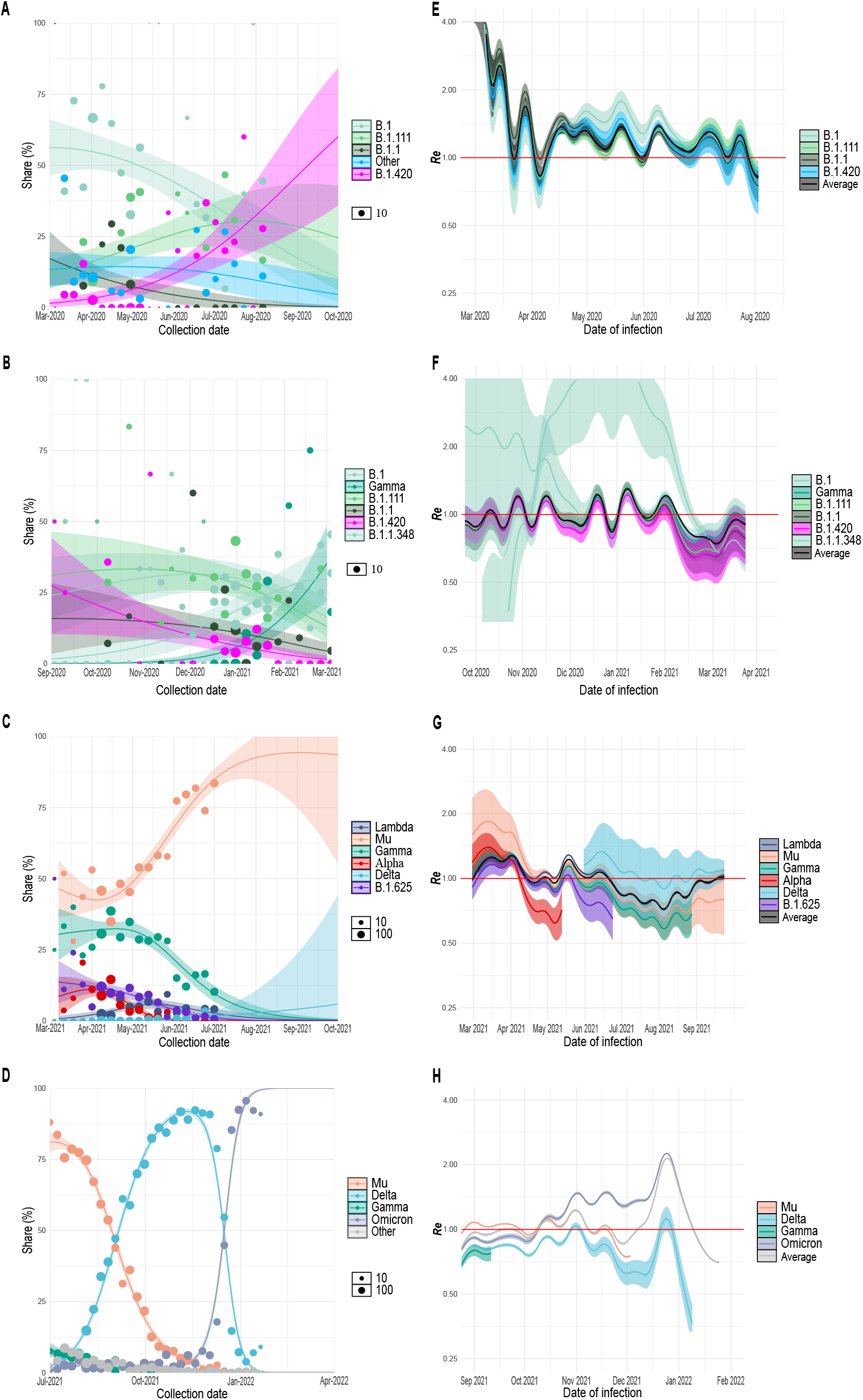
Multinomial fit analysis for the four periods (waves) of the SARS-CoV-2 pandemic in Colombia. A) to D) show a reconstruction of the growth advantage dynamics of the circulating variants during the four pandemic periods inferred from GISAID metadata90 whg+1and represented as share percentage. E) to H) show the *R*_*e*_ from each variant calculated from intrinsic growth rates as inferred from daily case data from WHO.

**Supplementary Fig.4.**
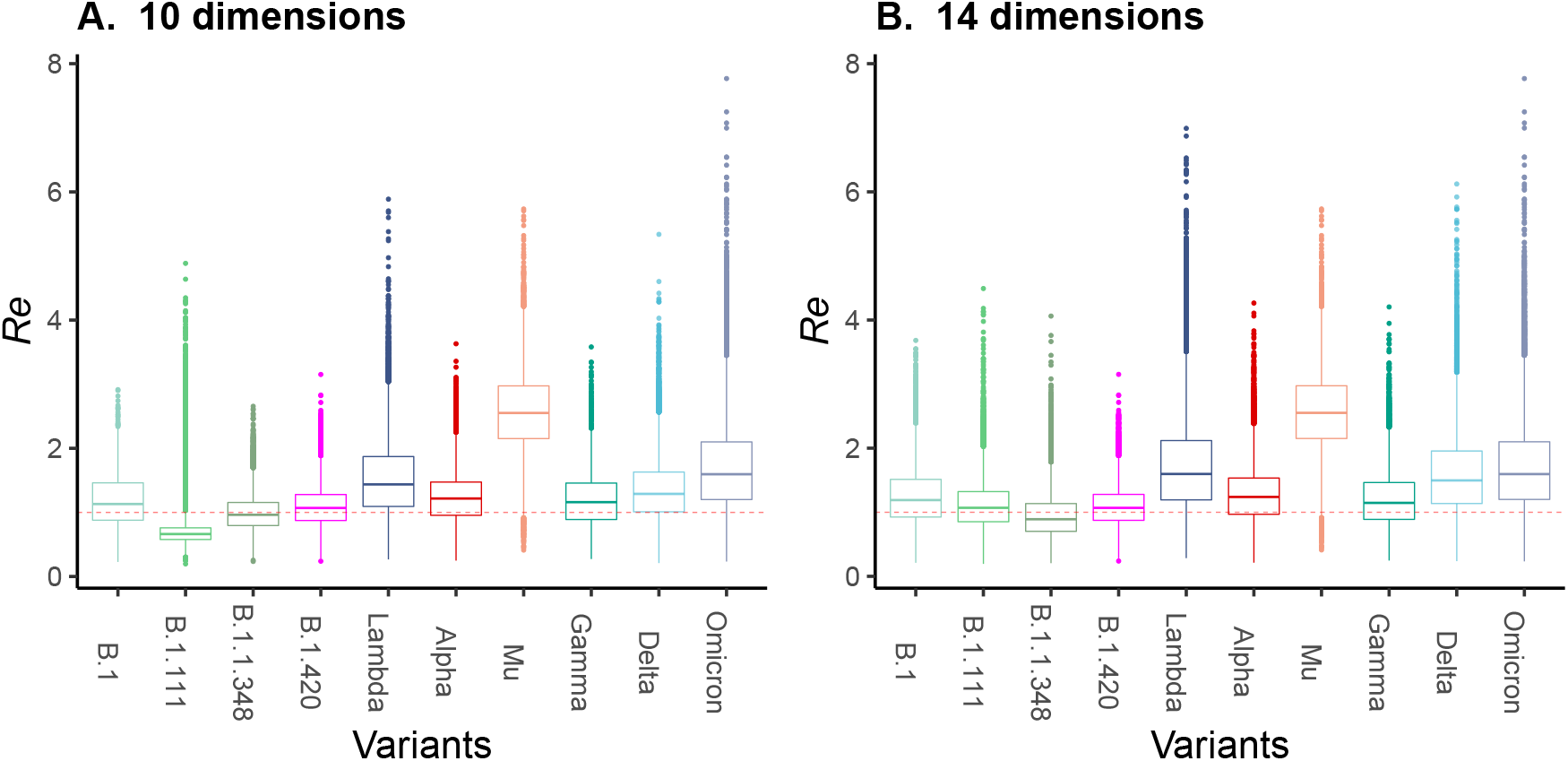
Global *R*_*e*_ estimations of the most prevalent variants (VOCIs). The effective reproduction number (*R*_*e*_) through time parameter was estimated using the Bayesian birth-death skyline model setting ten (A) and fourteen dimensions (B), showing congruence assuming different scenarios.

**Supplementary Fig.5.**
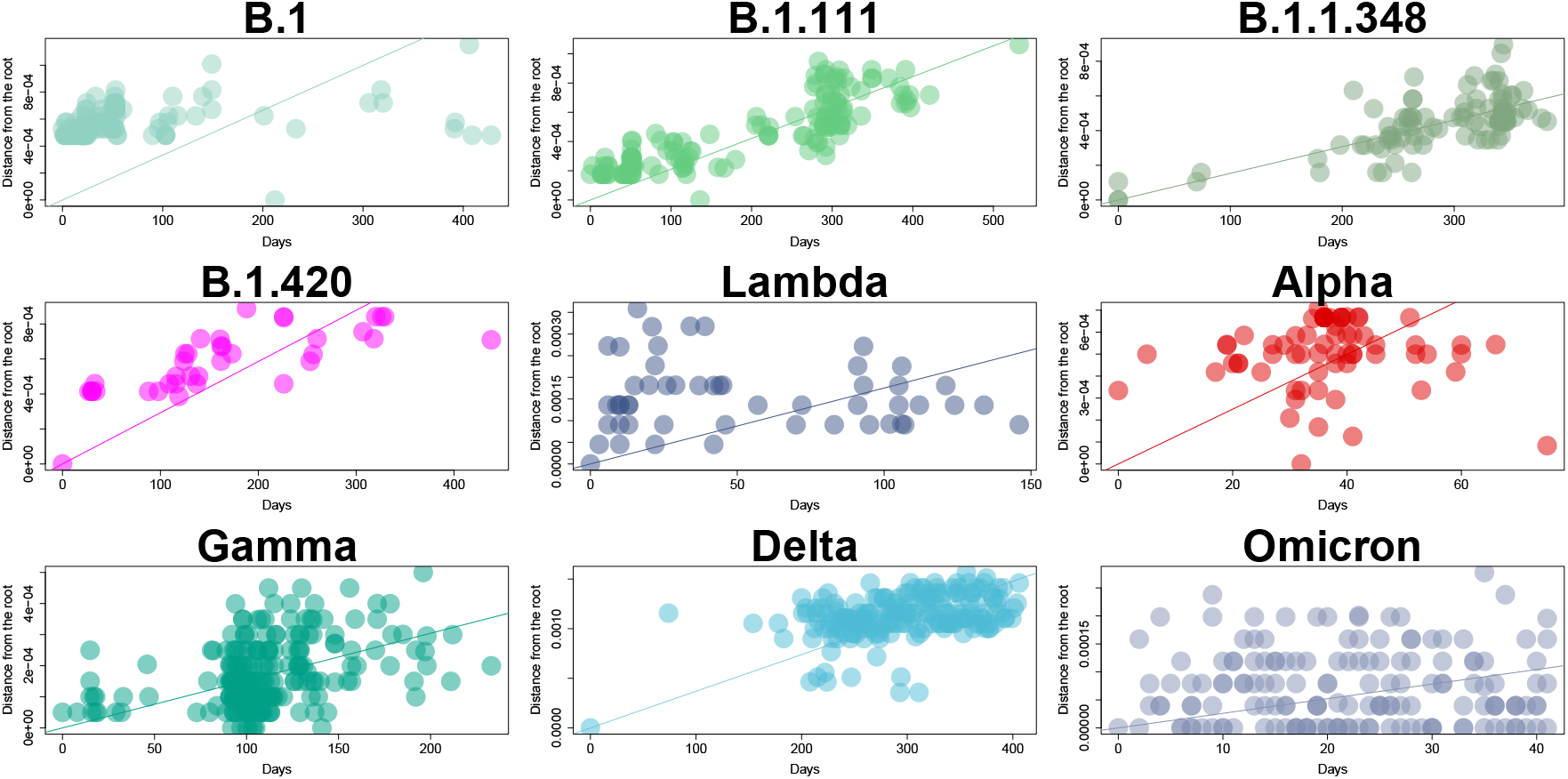
Root-to-tip regression analysis of Colombian SARS-CoV-2 sequences down-sampling alignments showing the phylogenetic signal per variant.

**Supplementary Fig.6.**
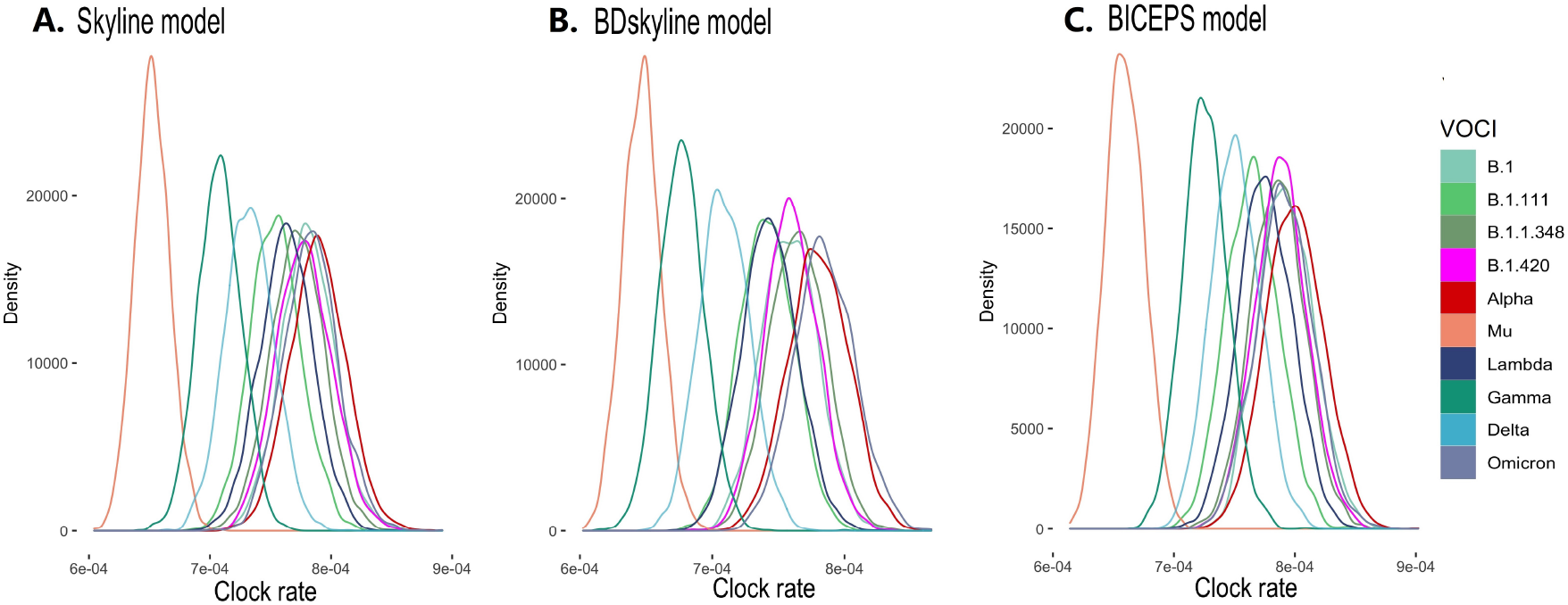
Density plot of the posterior of the Clock rate parameter for ten variants’ alignments (VOCIs). A) Coalescent Skyline model (Skyline), B) Birth-Death Skyline model (BDSkyline), C) Bayesian Integrated Coalescent Epoch PlotS (BICEPS).

## Notes

### Competing Interest Statement

The authors have declared no competing interest.

### Funding Statement

This work is supported by the Marsden grant 18-UOA-096 from the Royal Society of New Zealand, Colombia Science Ministry (Minciencias), BPIN 20200000100090. This project was funded by the Universidad del Rosario in the framework of its strategic plan RUTA2025. Thanks to President and the University Council for leading the strategic projects (JR). We also thank the Colombian network for SARS-CoV-2 genomic surveillance led by the National Institute of Health (INS).

### Author Declarations

Sample collection was led by the Instituto de Investigaciones Biologicas del Tropico at Universidad de Cordoba and the Centro de Investigaciones en Microbiologia y Biotecnologia at Universidad del Rosario, which are part of the official laboratories authorized by Colombias Ministry of Health for testing and genomic surveillance or SARS-CoV-2. Sample collection in Cordoba was approved by the Ethics committee of Universidad de Cordoba IIBT (Acta 0410-2020) in compliance with CDCs guidelines for safe work practices in human diagnostic. Sample collection in Bogota and Cali was approved by Universidad del Rosarios Research Ethics committee (DVO005 1550-CV1400) in compliance with Helsinkis declaration. Informed consent was obtained from all patients.

## References

1. Minsalud. Colombia confirma su primer caso de COVID-19 (2020).

2. Worldometer. COVID-19 coronavirus pandemic (2020). [Online on https://www.worldometers.info/coronavirus/; posted on 10 June 2020].

3. Hannah Ritchie, L. R.-G. C. A. C. G. E. O.-O. J. H. B. M. D. B., Edouard Mathieu & Roser, M. Coronavirus pandemic (covid-19). Our World Data (2020). Https://ourworldindata.org/coronavirus.

4. Castañeda, S. et al. Evolution and epidemic spread of sars-cov-2 in colombia: A year into the pandemic. Vaccines 9, 10.3390/vaccines9080837 (2021).

5. Flaxman, S. et al. Estimating the effects of non-pharmaceutical interventions on covid-19 in europe. Nature 584, 257–261 (2020).

6. Jaja, I. F., Anyanwu, M. U. & Jaja, C.-J. I. Social distancing: how religion, culture and burial ceremony undermine the effort to curb covid-19 in south africa. Emerg. Microbes & Infect. 9, 1077–1079, 10.1080/22221751.2020.1769501 (2020). PMID: 32459603, https://doi.org/10.1080/22221751.2020.1769501.

7. Dreher, N. et al. Policy interventions, social distancing, and sars-cov-2 transmission in the united states: a retrospective state-level analysis. The Am. journal medical sciences 361, 575–584 (2021).

8. Haug, N. et al. Ranking the effectiveness of worldwide COVID-19 government interventions. Nat. Hum. Behav. 2020 4:12 4, 1303–1312, 10.1038/s41562-020-01009-0 (2020).

9. Islam, S., Islam, T. & Islam, M. R. New coronavirus variants are creating more challenges to global healthcare system: a brief report on the current knowledge. Clin. Pathol. 15, 2632010X221075584 (2022).

10. Rambaut, A. et al. A dynamic nomenclature proposal for SARS-CoV-2 lineages to assist genomic epidemiology. Nat. Microbiol. 10.1038/s41564-020-0770-5 (2020).

11. Bernal, J. L. et al. Effectiveness of covid-19 vaccines against the b. 1.617. 2 (delta) variant. New Engl. J. Medicine (2021).

12. Li, L. et al. Transmission and containment of the sars-cov-2 delta variant of concern in guangzhou, china: A population-based study. PLoS neglected tropical diseases 16, e0010048 (2022).

13. Organization, W. H. Tracking sars-cov-2 variants.

14. Janik, E., Niemcewicz, M., Podogrocki, M., Majsterek, I. & Bijak, M. The emerging concern and interest sars-cov-2 variants, 10.3390/pathogens10060633 (2021).

15. Resende, P. C. et al. Evolutionary Dynamics and Dissemination Pattern of the SARS-CoV-2 Lineage B.1.1.33 During the Early Pandemic Phase in Brazil. Front. Microbiol. 11, 3565, 10.3389/fmicb.2020.615280 (2021).

16. Garcia-Beltran, W. F. et al. Multiple SARS-CoV-2 variants escape neutralization by vaccine-induced humoral immunity. Cell 184, 2372–2383.e9, 10.1016/j.cell.2021.03.013 (2021).

17. Volz, E. et al. Evaluating the Effects of SARS-CoV-2 Spike Mutation D614G on Transmissibility and Pathogenicity. Cell 184, 64–75.e11, 10.1016/j.cell.2020.11.020 (2021).

18. Konings, F. et al. SARS-CoV-2 Variants of Interest and Concern naming scheme conducive for global discourse, 10.1038/s41564-021-00932-w (2021).

19. Campbell, F. et al. Increased transmissibility and global spread of SARS-CoV-2 variants of concern as at June 2021. Eurosurveillance 26, 2100509, 10.2807/1560-7917.es.2021.26.24.2100509 (2021).

20. Laiton-Donato, K. et al. Characterization of the emerging B.1.621 variant of interest of SARS-CoV-2. Infect. Genet. Evol. 95, 105038, 10.1016/j.meegid.2021.105038 (2021).

21. Viana, R. et al. Rapid epidemic expansion of the SARS-CoV-2 Omicron variant in southern Africa. medRxiv 2021.12.19.21268028, 10.1101/2021.12.19.21268028 (2021).

22. Geoghegan, J. et al. Genomic epidemiology reveals transmission patterns and dynamics of sars-cov-2 in aotearoa new zealand, 10.1101/2020.08.05.20168930 (2020).

23. Douglas, J. et al. Phylodynamics reveals the role of human travel and contact tracing in controlling covid-19 in four island nations. medRxiv 10.1101/2020.08.04.20168518 (2020). https://www.medrxiv.org/content/early/2020/08/06/2020.08.04.20168518.full.pdf.

24. Instituto nacional de salud (ins).

25. Haug, N. et al. Ranking the effectiveness of worldwide covid-19 government interventions. Nat. human behaviour 4, 1303–1312 (2020).

26. Laiton-Donato, K. et al. Genomic epidemiology of sars-cov-2 in colombia, 10.1101/2020.06.26.20135715 (2020).

27. Ramírez, J. D. et al. The arrival and spread of SARS-CoV-2 in Colombia. J. Med. Virol. jmv.26393, 10.1002/jmv.26393 (2020).

28. Ministerio de Salud y Protección Social. Resolución 00000380 del Ministerio de Salud y Protección Social. 5 (2020).

29. Ministerio del Interior. Republica de Colombia. Decreto Número 457. Decreto Número 457 1–14 (2020).

30. Petrone, M. E. et al. Insights into the limited global spread of the immune evasive sars-cov-2 variant mu. medRxiv 2022–03 (2022).

31. Faria, N. R. et al. Genomics and epidemiology of the P.1 SARS-CoV-2 lineage in Manaus, Brazil. Science 372, 10.1126/science.abh2644 (2021).

32. du Plessis, L. et al. Establishment and lineage dynamics of the SARS-CoV-2 epidemic in the UK. Science 371, 708–712, 10.1126/SCIENCE.ABF2946/SUPPL_FILE/DUPLESSIS_SM.PDF (2021).

33. Naveca, F. G. et al. COVID-19 in Amazonas, Brazil, was driven by the persistence of endemic lineages and P.1 emergence. Nat. Medicine 27, 1230–1238, 10.1038/s41591-021-01378-7 (2021).

34. Uriu, K. et al. Neutralization of the SARS-CoV-2 Mu Variant by Convalescent and Vaccine Serum. New Engl. J. Medicine 385, 2397–2399, 10.1056/NEJMC2114706/SUPPL_FILE/NEJMC2114706_DISCLOSURES.PDF (2021).

35. Tada, T. et al. High-titer neutralization of Mu and C.1.2 SARS-CoV-2 variants by vaccine-elicited antibodies of previously infected individuals. Cell Reports 38, 110237, 10.1016/j.celrep.2021.110237 (2022).

36. Bushman, M., Kahn, R., Taylor, B. P., Lipsitch, M. & Hanage, W. P. Population impact of SARS-CoV-2 variants with enhanced transmissibility and/or partial immune escape. Cell 184, 6229–6242.e18, 10.1016/j.cell.2021.11.026 (2021).

37. Ramirez, A. L. et al. Impact of SARS-CoV-2 Mu variant on vaccine effectiveness: A comparative genomics study at the peak of the third wave in Bogota, Colombia. J. Med. Virol. 10.1002/JMV.27808 (2022).

38. Uriu, K. et al. Characterization of the Immune Resistance of Severe Acute Respiratory Syndrome Coronavirus 2 Mu Variant and the Robust Immunity Induced by Mu Infection. The J. Infect. Dis. 10.1093/infdis/jiac053 (2022).

39. Liu, Y. & Rocklöv, J. The reproductive number of the Delta variant of SARS-CoV-2 is far higher compared to the ancestral SARS-CoV-2 virus. J. travel medicine 28, 10.1093/jtm/taab124 (2021).

40. Cai, J. et al. Modeling transmission of SARS-CoV-2 Omicron in China. Nat. medicine 1–1, 10.1038/s41591-022-01855-7 (2022).

41. Serrano-Coll, H. et al. Effectiveness of the CoronaVac® vaccine in a region of the Colombian Amazon, was herd immunity achieved? Trop. Dis. Travel. Medicine Vaccines 8, 4–9, 10.1186/s40794-021-00159-x (2022).

42. Mattar, S. et al. Severe Acute Respiratory Syndrome Coronavirus 2 Seroprevalence among Adults in a Tropical City of the Caribbean Area, Colombia: Are We Much Closer to Herd Immunity Than Developed Countries? Open Forum Infect. Dis. 7, 10.1093/ofid/ofaa550 (2020).

43. Laajaj, R. et al. COVID-19 spread, detection, and dynamics in Bogota, Colombia. Nat. Commun. 12, 10.1038/s41467-021-25038-z (2021).

44. Douglas, J. et al. Real-time genomics for tracking severe acute respiratory syndrome coronavirus 2 border incursions after virus elimination, new zealand. Emerg. Infect. Dis. 27, 2361 (2021).

45. Gu, H. et al. Genomic epidemiology of SARS-CoV-2 under an elimination strategy in Hong Kong. Nat. Commun. 13, 10.1038/s41467-022-28420-7 (2022).

46. Bouckaert, R. R. An efficient coalescent epoch model for bayesian phylogenetic inference. bioRxiv 10.1101/2021.06.28.450225 (2021). https://www.biorxiv.org/content/early/2021/06/30/2021.06.28.450225.full.pdf.

47. Shu, Y. & McCauley, J. GISAID: Global initiative on sharing all influenza data – from vision to reality. Euro. Surveill. 22, 30494 (2017).

48. Hadfield, J. et al. Nextstrain: real-time tracking of pathogen evolution. Bioinformatics 34, 4121–4123 (2018).

49. Argimón, S. et al. Microreact: visualizing and sharing data for genomic epidemiology and phylogeography. Microb. genomics 2 (2016).

50. Corman, V. M. et al. Detection of 2019 novel coronavirus (2019-nCoV) by real-time RT-PCR. Euro surveillance : bulletin Eur. sur les maladies transmissibles = Eur. communicable disease bulletin 25, 1–8, 10.2807/1560-7917.ES.2020.25.3.2000045 (2020).

51. Quick, J. ncov-2019 sequencing protocol v3 (locost) v.3.

52. Wick, R. R., Judd, L. M. & Holt, K. E. Performance of neural network basecalling tools for oxford nanopore sequencing. Genome biology 20, 1–10 (2019).

53. Nick Loman, A. R., Will Rowe. ncov-2019 sequencing protocol v3 (locost) v.3.

54. Miller, J. M. et al. Guidelines for Safe Work Practices in Human and Animal Medical Diagnostic Laboratories Recommendations of a CDC-convened, Biosafety Blue Ribbon Panel Centers for Disease Control and Prevention MMWR Editorial and Production Staff MMWR Editorial Board. Centers for Dis. Control. Prev. Morb. Mortal. Wkly. Rep. 61, 105 (2012).

55. World Medical Association Declaration of Helsinki: ethical principles for medical research involving human subjects. The J. Am. Coll. Dent. 81, 14–18, 10.1093/acprof:oso/9780199241323.003.0025 (2014).

56. O’Toole, Á. et al. Assignment of epidemiological lineages in an emerging pandemic using the pangolin tool. Virus Evol. 7, 10.1093/ve/veab064 (2021).

57. Aksamentov, I., Roemer, C., Hodcroft, E. B. & Neher, R. A. Nextclade: clade assignment, mutation calling and quality control for viral genomes. J. Open Source Softw. 6, 3773, 10.21105/joss.03773 (2021).

58. Hadfield, J. et al. Nextstrain: real-time tracking of pathogen evolution. Bioinformatics 34, 4121–4123, 10.1093/bioinformatics/bty407 (2018). https://academic.oup.com/bioinformatics/article-pdf/34/23/4121/26676762/bty407.pdf.

59. R Core Team. R: A Language and Environment for Statistical Computing. R Foundation for Statistical Computing, Vienna, Austria (2014).

60. Davies, N. G. et al. Increased mortality in community-tested cases of SARS-CoV-2 lineage B.1.1.7. Nat. 2021 593:7858 593, 270–274, 10.1038/s41586-021-03426-1 (2021).

61. Nishiura, H., Linton, N. M. & Akhmetzhanov, A. R. Serial interval of novel coronavirus (covid-19) infections. International Journal of Infectious Diseases (2020).

62. Katoh, K., Asimenos, G. & Toh, H. Multiple alignment of DNA sequences with MAFFT. In Bioinformatics for DNA sequence analysis, 39–64 (Springer, 2009).

63. Paradis, E. et al. Package ‘ape’. Analyses phylogenetics evolution, version 2 (2019).

64. Rambaut, A., Lam, T. T., Max Carvalho, L. & Pybus, O. G. Exploring the temporal structure of heterochronous sequences using tempest (formerly path-o-gen). Virus evolution 2, vew007 (2016).

65. Drummond, A. J., Rambaut, A., Shapiro, B. & Pybus, O. G. Bayesian coalescent inference of past population dynamics from molecular sequences. Mol. biology evolution 22, 1185–1192 (2005).

66. Stadler, T., Kühnert, D., Bonhoeffer, S. & Drummond, A. J. Birth–death skyline plot reveals temporal changes of epidemic spread in hiv and hepatitis c virus (hcv). Proc. Natl. Acad. Sci. 110, 228–233 (2013).

67. Stadler, T., Kühnert, D., Bonhoeffer, S. & Drummond, A. J. Birth–death skyline plot reveals temporal changes of epidemic spread in hiv and hepatitis c virus (hcv). Proc. Natl. Acad. Sci. 110, 228–233, 10.1073/pnas.1207965110 (2013). https://www.pnas.org/content/110/1/228.full.pdf.

68. Lemey, P., Rambaut, A., Drummond, A. J. & Suchard, M. A. Bayesian phylogeography finds its roots. PLoS Comp. Biol. 5 (2009).

69. Hasegawa, M., Kishino, H. & aki Yano, T. Dating of the human-ape splitting by a molecular clock of mitochondrial dna. J. Mol. Evol. 22, 160–174 (1985).

70. Bouckaert, R. R. & Drummond, A. J. bModelTest: Bayesian phylogenetic site model averaging and model comparison. BMC Evol. Biol. 17, 42 (2017).

71. Müller, N. F. & Bouckaert, R. Coupled MCMC in BEAST 2. bioRxiv (2019).

72. Paradis, E., Claude, J. & Strimmer, K. APE: Analyses of Phylogenetics and Evolution in R language. Bioinformatics 20, 289–290, 10.1093/bioinformatics/btg412 (2004).

73. Yu, G. Using ggtree to visualize data on tree-like structures. Curr. Protoc. Bioinforma. 69, e96, https://doi.org/10.1002/cpbi.96 (2020). https://currentprotocols.onlinelibrary.wiley.com/doi/pdf/10.1002/cpbi.96.

74. for Health Metrics, I. & (IHME), E. Covid-19 projections.

75. (OxCGRT), O. C.-. G. R. T. Covid-19 government response tracker.

